# Phase locking of auditory steady state responses is modulated by a predictive sensory context and linked to degree of myelination in the cerebellum

**DOI:** 10.1101/2023.06.08.23291140

**Authors:** Kit Melissa Larsen, Kiran Thapaliya, Markus Barth, Hartwig R. Siebner, Marta I. Garrido

**Author notes:** Corresponding author: Melissa Larsen.

## Abstract

**Background:** 40 Hz auditory steady state responses (ASSR) can be evoked by brief auditory clicks delivered at 40 Hz. While the neuropharmacology behind the generation of ASSR is well examined, the link between ASSR and microstructural properties of the brain is unclear. Further, whether the 40 Hz ASSR can be manipulated through processes involving top-down control, such as prediction, is currently unknown.

**Methods:** We recorded EEG in 50 neurotypical participants while they engaged in a 40 Hz Auditory steady state paradigm. We manipulated the predictability of tones to test the modulatory effect of prediction on 40 Hz steady state responses. Further, we acquired T1w and T2w structural MRI and used the T1/T2 ratio as a proxy to determine myelination in grey matter.

**Results:** The phase locking of the 40 Hz ASSR was indeed modulated by prediction and this modulation extended to all frequency bands, suggesting prediction violation as a phase resetting mechanism. Interestingly, we found that the prediction violation of the phase locking at 40 Hz (gamma) was associated with the degree of grey matter myelination in the right cerebellum.

**Discussion:** We demonstrate that prediction violations evoke resetting of oscillatory activity and suggest that the efficiency of this process is promoted by greater cerebellar myelin. Our findings provide a structural-functional relationship for myelin and phase locking of auditory oscillatory activity. These results introduce a setting for looking at the interaction of predictive processes and ASSR in disorders where these processes are impaired such as in psychosis.

## Introduction

The generation of synchronous activity in the brain underpins coordinated activity and connectivity among functional networks. Consistent evidence shows that theta and gamma oscillations propagate via feedforward connections whereas beta and alpha oscillations propagate through backward connections (Bastos et al., 2015; Jensen et al., 2015; Kerkoerle et al., 2014). Given the role of oscillatory activity in enabling brain connectivity, the interplay between frequency bands may therefore enable interplay between bottom-up and top-down processing. In fact, in the visual domain, beta activity enhances gamma activity (Richter et al., 2017) indicating an interplay between top-down and bottom-up processing. In this way top-down control can enhance the ability to generate gamma oscillations. One way to evoke cortical gamma oscillations is through exposure to auditory stimuli by brief tones or clicks at repetition rates of 40Hz. In this way, an auditory steady state response (ASSR) is elicited and can be measured with electroencephalography (EEG). These cortical gamma oscillations rely on the integrity of fast-spiking GABAergic interneurons which exert a finely timed inhibition onto the pyramidal cells and other inhibitory interneurons (Bartos et al., 2007; Sohal et al., 2009; Traub et al., 2003). Both the phase and power of the ASSR are believed to reflect the inhibitory/excitatory balance mediated by the N-methyl-D-aspartate (NMDA) receptor (Sivarao et al., 2016; Tada et al., 2020). Whether the 40 Hz ASSR can be manipulated through processes involving top-down control, such as predictive processing, is currently unknown. The ability to make predictions about the sensory environment is a crucial feature of the brain (Friston, 2005). Detecting violations to those predictions such as changes in the acoustic environment is critical for survival as these indicate potential threats or rewards. A classic way to investigate this process in the lab is to use a sequency of auditory streams of highly probable standard sounds and surprising deviant sounds that generates a mismatch response (Fitzgerald and Todd, 2020).

While neuropharmacological studies have linked ASSR to NMDA and GABA (Sivarao, 2015; Sohal et al., 2009; Tada et al., 2020), the relationship between the expression of ASSR and microstructural properties of the brain is unclear. T1-weighted (T1w) and T2-weighted (T2w) images can be combined to assess tissue microstructure via T1w/T2w ratio maps (Ganzetti et al., 2014). Whole brain T1w/T2w ratio maps have provided a sensitive measure, related to myelin and microstructural integrity in conditions such as multiple sclerosis (Beer et al., 2016) and schizophrenia (Ganzetti et al., 2015). Recently, grey matter myelin has been linked to electrophysiological connectivity (Hunt et al., 2016), with the strongest relations to the beta and gamma band. Similarly, the rhythmicity of high frequency oscillations is related to cortical myelin content (Tomasevic et al., 2022) and activity-dependent myelination have shown to promote neural phase synchronization (Noori et al., 2020). Hence, there is increasing evidence that the microstructure of the cortex supports functional networks. The idea of myelin playing a role in the ability to generate oscillatory synchrony becomes clear when noting that myelin speeds up conduction velocity (Dutta et al., 2018). In this way even small changes in the conduction velocity can impact the generation of synchrony and coupling between brain regions (Pajevic et al., 2014). A link between the functional ASSR and structural atrophy in the auditory cortex has been established (Mancini et al., 2022). Though, it is still unknown if the power and phase locking of the ASSR scale with the degree of cortical myelination.

Disturbed interactions of the circuits underlying ASSR are thought to critically contribute to pathogenesis and cognitive impairment in neurodevelopmental disorders such as schizophrenia (Gonzalez-Burgos et al., 2011; Jadi et al., 2016; Thuné et al., 2016). Reduction in the 40 Hz ASSR is not only seen in chronic schizophrenia, but also in first episode psychosis (Spencer et al., 2008; Symond et al., 2005), as well as in non-affected first-degree relatives (Hong et al., 2004; Rass et al., 2012). Given that both predictive processes and 40 Hz ASSR are reduced in psychosis, introducing ASSR in a predictive setting will provide insights into how low-level auditory processing and higher-level network integration may go awry in these disorders. Mounting evidence suggesting that psychotic-like experiences occur to a certain extent in the healthy general population (Verdoux and van Os, 2002), have led to the formulation of the continuum of psychosis hypothesis. According to this hypothesis every individual in the general population lies somewhere on a continuum of psychotic-like traits, with some individuals scoring high on the personality dimension of schizotypy, i.e., psychotic-like experiences. Indeed, some of the behavioral, functional, and electrophysiological deficits seen in clinical psychosis, are also observed to some extent in the healthy population see for example (Dzafic et al., 2021; Jahshan et al., 2012; Oestreich et al., 2019). However, how ASSR scales with schizotypal personality traits in individuals without a disorder, is unknown.

Given the link between reduced ASSR and neurodevelopmental disorders, here we ask whether psychotic-like experiences in the general population are associated with inter-individual differences in ASSR. We introduce ASSR in a predictive setting integrating low level auditory processing with higher level network processes. We ask whether 40 Hz ASSR can be manipulated by predictability and whether these electrophysiological readouts are related to the degree of myelination.

## Methods

### Participants

50 healthy adults 18-25 years were recruited through the Psychology Research Participation Scheme (SONA) at the University of Queensland. Prior screening ruled out participants reporting a history of psychiatric or neurological disorders and currently on medication acting on the central nervous system. Participants completed the 92 Item-Prodromal questionnaire (PQ), which measures positive and negative symptoms and is typically used to assess psychotic experiences in healthy individuals (L.Loewy et al., 2005). In addition, participants completed the Beck Depression questionnaire (Beck et al., 1988b) and Beck anxiety (Beck et al., 1988a). To ensure that the PQ scores was not mainly driven by depressive or anxiety symptoms, participants meeting the threshold for moderate anxiety or depression were excluded. Participants provided written informed consent and received monetary reimbursement for their time. This research was approved by the University of Queensland Human Research Ethics.

### Paradigm

Participants were presented with a 40 Hz auditory steady state paradigm with the 40 Hz click trains arranged in a traditional oddball paradigm, while we measured EEG (see Figure 1). In this way it was possible to evoke 40 Hz gamma oscillations through click trains with the manipulation of expectancy of the tone. The design was divided into two blocks where deviant tones were longer than the standards in the long block and vice versa in the short block. This allowed us to compare the deviant tones from the long block to the standard tones of the short block ensuring the comparison of tones with the same physical properties but with a change in expectancy of the tone duration (even though duration was matched). We divided each block into two runs of approximately 8 minutes, meaning that the total recording time was (4×8minutes). This ensured that we had at least 180 trials for the deviant tones in each block. Runs and blocks were counterbalanced across subjects. To keep participants engaged, they performed a visual 1-back task, which entailed detecting repetitions of letters continuously presented, which did not coincide with the sounds.

**Figure 1:**
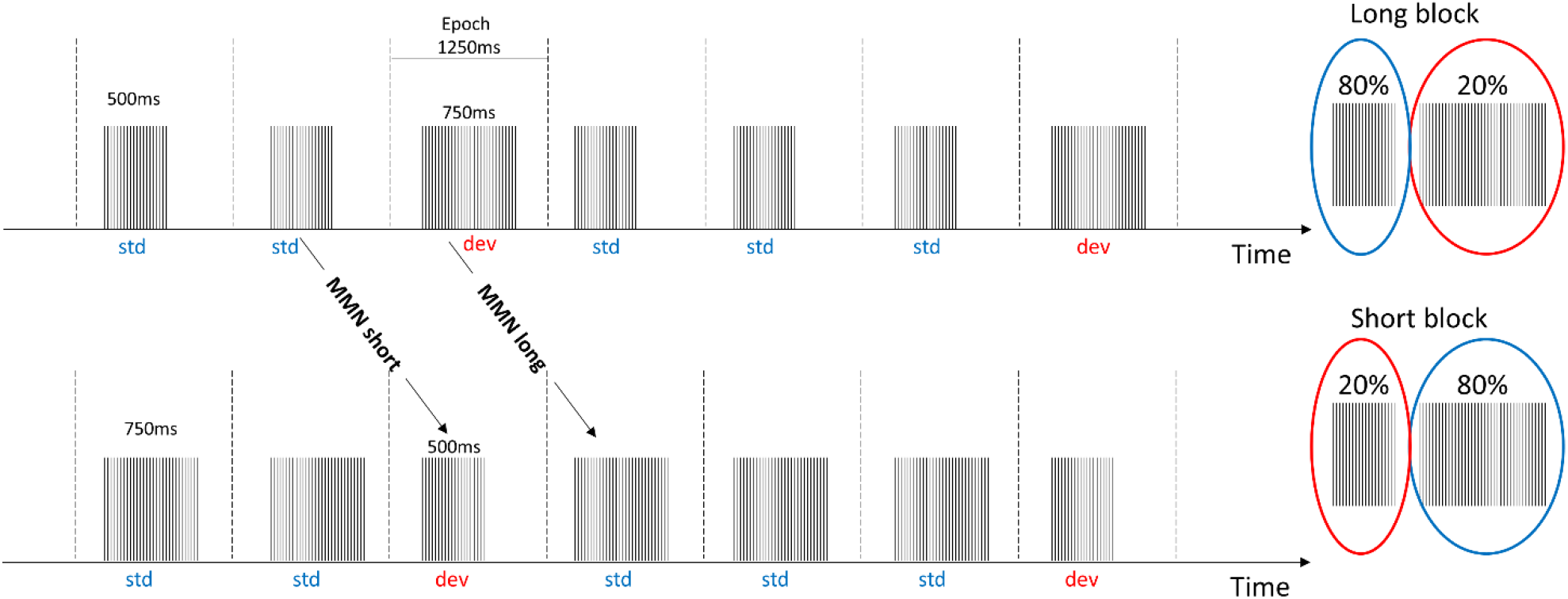
Click trains with a frequency of 40 Hz were arranged in a classical oddball paradigm where the deviant changed tone duration to either be longer than the standard (long block) or shorter than the standard (short block). The MMN_short_ responses were derived by contrasting the standard tone in the long block with the deviant tone in the short block, in order to ensure comparison of tones with the same duration. Likewise, the MMN_long_ were derived by contrasting the standard tone in the short block with the deviant tones in the long block. Epochs were extracted from 250 ms pre-stimulus to 1000 ms post-stimulus onset.

### EEG and preprocessing

EEG data was recorded using a 64 channel Biosemi active two with electrodes arranged according to the 10-20 system and a sampling frequency of 1024 Hz. All offline preprocessing was performed using SPM 12 (http://www.fil.ion.ucl.ac.uk/spm/), which included high and low pass filtering with a 5^th^ order Butterworth filter with a cut-off of 0.5 Hz and 80 Hz respectively. A notch filter was applied with cutoff 48-52 Hz. Data were epoched with a peristimulus interval of -250 ms to 1000 ms, with baseline correction applied from -250ms to 0 ms. Finally, artefact rejection was performed using a simple threshold technique rejecting trials with amplitudes exceeding ±100 μV. Finally, the signals were referenced to the average of the two mastoid electrodes.

### Time-frequency analysis

The epoched data were wavelet-transformed using a Complex Morlet wavelet with 7 cycles and frequency band of interest covering frequencies from 2 to 80 Hz at 1 Hz resolution. Power and phase locking factor (PLF) were extracted from the wavelet coefficients. The amplitude of the PLF reflects the phase consistency across trials for a given channel, time, and frequency point. Values of PLF are bounded between 0 to 1, with 1 being perfect phase synchrony across trials. To compare power and PLF values for the different frequency bands, values were extracted during the time of stimulation 150-500 ms and for the standard frequency bands of theta (4-7 Hz), alpha (8-12 Hz), beta (15-25 Hz) and gamma (35-45Hz). We analyse the whole data spectrum and show the spatial distribution of the effects on topographical plots. We then zoom in on data extracted from electrode Cz where 40 Hz ASSR typically show the largest amplitude.

### Approximation of level of myelination

To get an approximation for myelin-content a T1 weighted magnetization prepared rapid gradient-echo (MPRAGE) and a T2 SPACE sequence were acquired for all participants on a Siemens 3T Prisma scanner with a 64-channel head coil. T1 weighted data were acquired using the following parameters: repetition time (TR)=4s, echo time (TE) =29ms, flip angle (FA) = 6^0^and an isotropic resolution of 1mm with matrix size = [176 240 256].

Similarly, T2 weighted data were acquired using TR=3.2s, TE=0.408s, TR=3.2, FA=120^0^, resolution of1mm isotropic with matrix size = [176 256 256]. The T1w/T2w ratio maps were calculated using the MRtool integrated in SPM12 (Ganzetti et al., 2015, 2014). Pre-processing steps include bias correction, intensity normalization, and computation of T1w/T2w ratio in the MNI space. We segmented T1w images into grey and white matter using the segmentation implemented in SPM12. Based on this segmentation we extracted T1w/T2w value of the grey matter regions. We focused on the grey matter since this generates the ASSR (Farahani et al., 2021). Before using these in the GLMs, images were smoothed using a 4×4×4 mm kernel.

### Relationship between EEG measures and degree of myelination

To investigate the relationship between the power and phase-locking factor (PLF) of the different frequency bands of the EEG evoked ASSR, we performed separate GLMs with the T1w/T2w ratio images as dependent variable. One GLM per measure (power and PLF) for the grey matter were performed with the EEG measure for each frequency band as regressors and age and gender as covariates. We further included total grey matter volume as a covariate to control for individual volume levels. All results are reported for surviving clusters corrected at p<0.05 FWE, with a cluster forming threshold of p<0.001 uncorrected. For significant clusters we extracted the values of the T1w/T2w images in a 10mm square around the peak coordinate of the cluster. This extraction was performed to visualize the intensity values and the EEG measures.

## Results

Three participants were excluded due to scores reaching thresholds for severe anxiety or moderate depression on the Beck’s inventories. Additionally, two participants were excluded due to very noisy EEG data, resulting in 45 participants in the main analyses (mean age 21.32 ± 1.97, 27 females, 18 males). PQ scores were distributed between 5 and128 with a mean of 55.6 ± 31.5.

### Phase locking across trials is modulated by prediction

To test if predictability can modulate power and phase locking across trials, we compared the responses for standard and deviant sounds in the gamma band (see upper row of Figure 2A for power and lower row for PLF). Indeed, phase locking to the stimulus across trials was modulated by sound predictability in the gamma band, indicated by an increase in phase locking factor for deviant trials compared to standard trials (see time frequency plot in Figure 2 for the long blocks). Overall, there were no difference between long and short blocks, therefore, the figures show the long block and we have pulled the data together when reporting the statistical findings.

**Figure 2:**
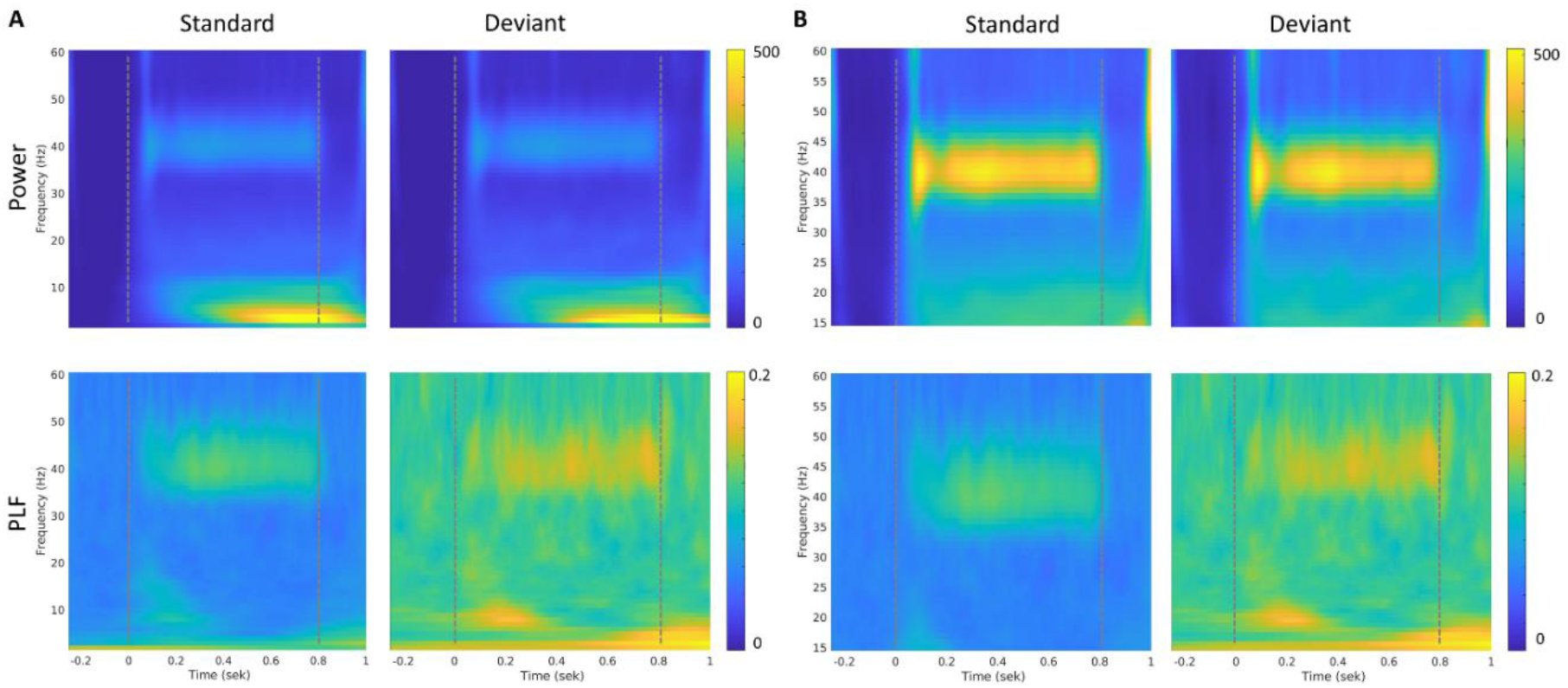
Time-frequency responses at channel Cz averaged across participants (for the long block, similar findings in the short block, see sup Figure 2). Dotted lines indicate onset and offset of auditory stimuli. A) Full frequency spectrum covering 2-60 Hz. B) is zoomed in at 15-60Hz to highlight the successful evoking of 40 Hz. All figures with power values have the same scale 0-500 and similarly for figures with phase locking factors, the scale is kept constant at 0 – 0.2.

Extracting values of power and phase locking factor, showed a robust difference between the standard and deviant trials, consistent across participants, (see Figure 3) with extracted values from channel Cz (where peak amplitudes for the 40 Hz ASSR is usually observed, see topography in supplementary Figure 1). In fact, this increase in phase locking factor for deviant trials was consistent across frequency bands, whereas power was similar for standard and deviant trials across all bands except for theta, where it showed a slight decrease in power for deviant trials compared to standard trials. For a topographic representation of the different frequency bands, (see supplementary Figure 1). To check that our results across frequency bands were not due to spillover effects, we repeated our analyses, this time filtering the data into the different frequency bands. Extracting the frequency values using this approach, did not change the results.

**Figure 3:**
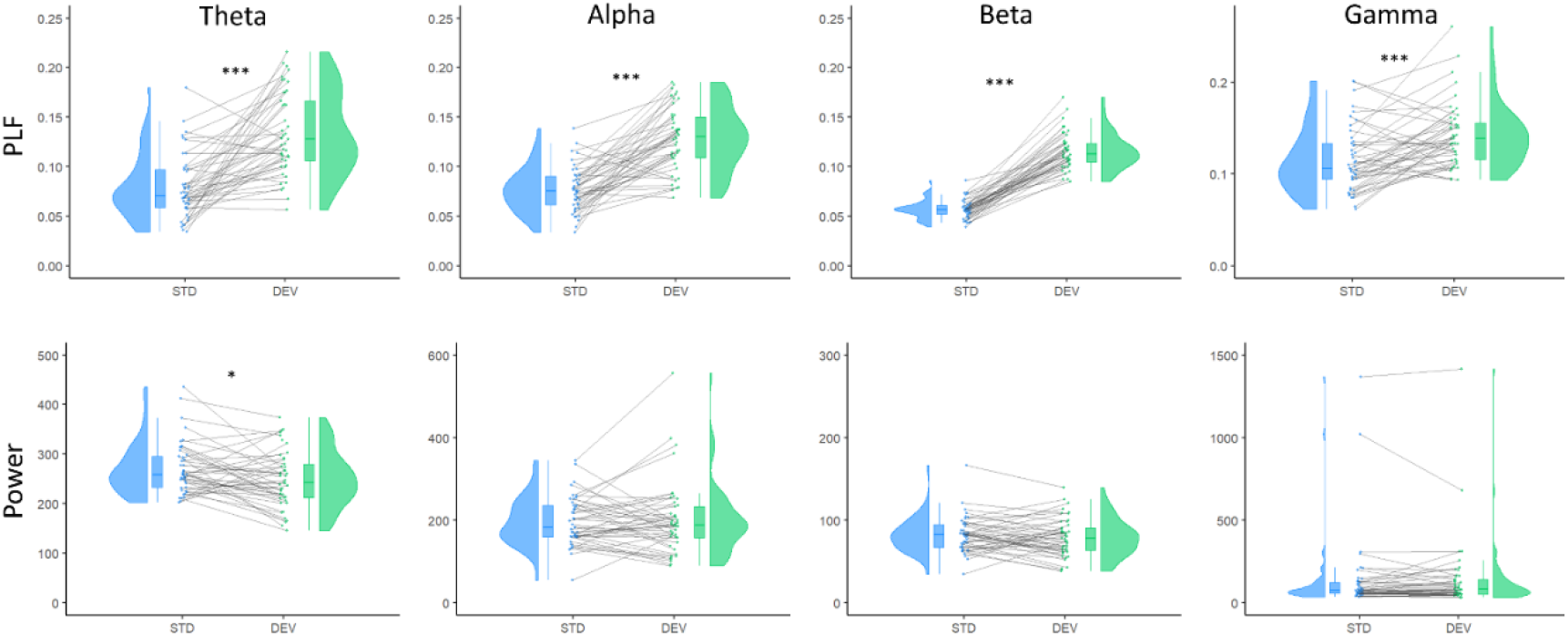
Power (bottom) and phase locking factor (top) extracted from channel Cz in the period of stimulation averaged across the long and short blocks. Blue indicates standard and green deviant. The connecting grey line indicates the relationship between the standard and deviant for each individual participant. * is p<0.05, *** is p < 0.001.

### The ability to phase lock to 40 Hz ASSR increases with greater myelin content

Since ASSR is generated in the grey matter, we performed a GLM using the grey matter T1w/T2w intensity images (Farahani et al., 2021). The difference in phase locking factor between the standard and deviant were positively associated with myelin content in the right cerebellum (Crus I), with peak values in [MNI 53 -57 -34] (see Figure 4A). Figure 4B shows the extracted T1w/T2w intensities in the significant cluster together with the phase locking values (difference between standard and deviant).

**Figure 4:**
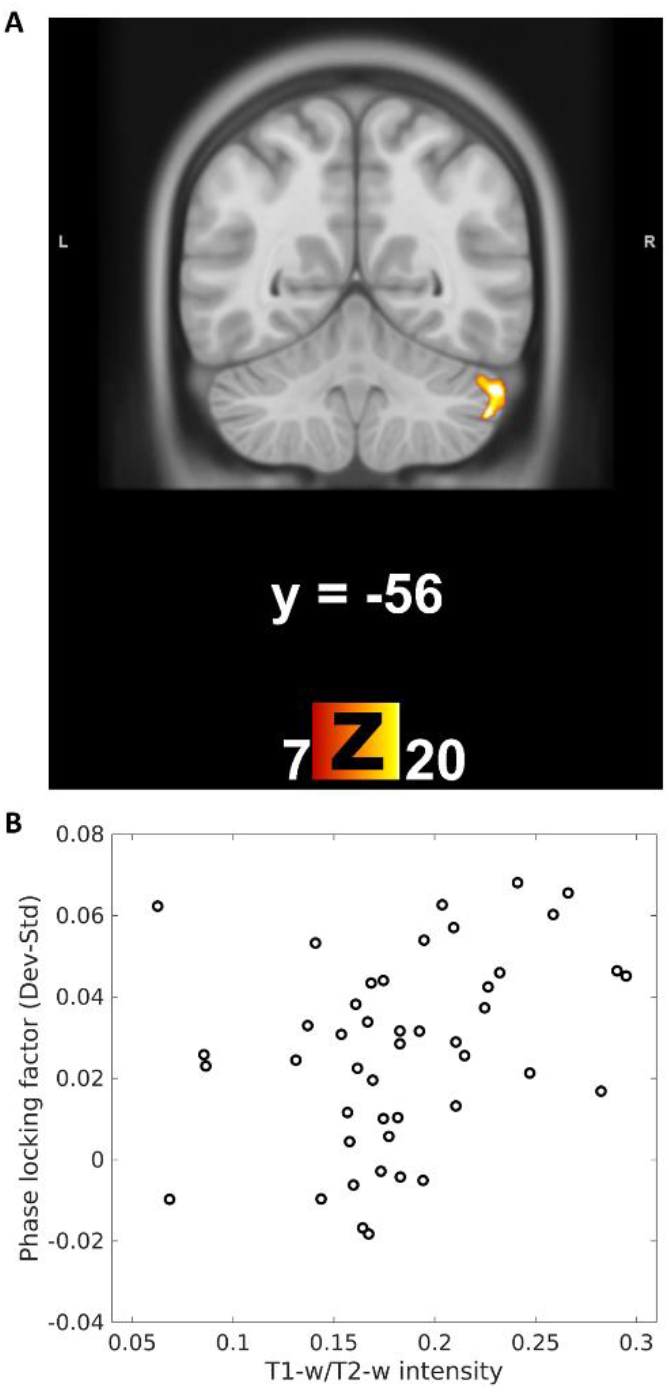
A: Association between myelin content and phase-locking for the gamma band (between the standard and deviant). Peak of the association was found in MNI coordinates [53 -57 -34]. Cluster forming threshold was set to p=0.001 uncorrected and results here only show the cluster that survived correction at the cluster level at p<0.05. B: T1w/T2w intensity values extracted from the cluster in the cerebellum increase with the Phase locking factor of the difference wave (between the standards and deviants).

### No evidence that 40 Hz ASSR is correlated with psychotic-like experiences

There was no correlation between the amount of gamma oscillation (both power and phase locking factor) and the degree of psychotic-like experiences. Given that prediction violation was not specific to the gamma band but modulated phase-locking across all four frequency bands, we post-hoc investigated the relationship between PLF for the theta, alpha and beta bands and PQ scores. However, no correlations were significant.

## Discussion

Here, we show that the ability to generate synchronous activity in the 40 Hz range through ASSRs, can be modulated by prediction. This modulation of prediction on the phase locking of the ASSR were linked to the myelin content in the grey matter of the cerebellum, potentially suggesting a mechanism for the ability to generate synchronous neural activity. Contrary to our hypothesis, we did not find any association between gamma range and degree of psychotic like experiences.

### ASSR is modulated by prediction

The violation of prediction induced by the oddball paradigm modulated not only the synchronization to the 40 Hz stimulation, but showed a modulation across all four frequency bands, as indicated by higher phase locking values for deviant responses compared to standard responses. This suggests that modulation is broadband rather than specific to the frequency of stimulation (40 Hz), and speaks to an overall deviant induced phase resetting mechanism (Canavier, 2015). The exact mechanisms behind phase resetting are unknown. However, the timing of ongoing oscillations can be altered through attentional processes (Kayser, 2009). Whether the increased phase locking for the deviant stimuli across frequency bands comes from directed attention towards the deviant or whether it is the prediction violation itself cannot be disentangled in the present study. As mentioned, prediction increased the phase locking factor across all frequency bands, but this increase was most consistent in the beta band, as the effect was present in every participant. Activity in the beta band is involved in predictive processes (Betti et al., 2021; van Pelt et al., 2016) and is believed to drive activity at 40 Hz (Metzner and Steuber, 2021). In the visual domain, beta oscillations enhance gamma activity (Richter, Thompson, Bosman, & Fries, 2017). Since beta and gamma have been associated with top-down and bottom-up processes respectively, our results are in keeping with the idea that top-down control can enhance bottom-up processes (Richter 2017). In a similar vein, our results suggest that beta may also have a role in driving 40 Hz synchronization in the auditory system.

### Structure-function relationship

We asked whether the ability to generate oscillatory synchrony is associated with the degree of myelination. We indeed found that phase locking to the auditory stimulation at 40 Hz were associated with cortical myelination in the right cerebellum, more specifically the right Crus I. While the cortical generator of the ASSR is typically seen spreading over the auditory areas (temporal regions) (Arutiunian et al., 2022; Farahani et al., 2021) and cerebellum (Pastor et al., 2002), the frontal areas are also involved in the mismatch negativity responses (Garrido et al., 2009). Our results therefore suggest that within the same areas generating the 40 Hz ASSR, the ability to phase lock to the 40 Hz auditory stimulation is associated with the degree of myelination within those regions. Interestingly, we showed that the association between oscillatory activity and cortical myelination were only present for the phase locking and not magnitude of the oscillatory activity. Similar results have recently been found using somatosensory stimulation (Tomasevic et al., 2022). They showed that the rhythmicity of high frequency oscillations within the human sensorimotor cortex was related to the cortical myelin content in the primary sensory area. Consistent with our findings, this association was only found for the rhythmicity and not the magnitude of the frequency content. Given the role of myelin in conducting velocity, even small changes in myelin can cause changes to the phase of the signal propagation (Pajevic et al., 2014).

While the T1w/T2w map is sensitive to the cortical myelin content (Ganzetti et al., 2014; Glasser and van Essen, 2011) it is not a direct quantitative measure of myelin. This is a limitation when comparing between studies with different scanning parameters, which can have an impact on the approximation of myelin content.

### No evidence that 40Hz ASSR is correlated with PQ

Inspired by the idea of the continuum of psychosis (van Os et al., 2009), we hypothesized that the 40Hz ASSR would scale with the degree of psychotic like experiences. However, our data did not support this hypothesis. While 40 Hz ASSR is reduced in first degree relatives (Hong et al., 2004; Rass et al., 2012; Thuné et al., 2016) and in individuals at clinical (Grent-’t-Jong et al., 2021) or genetic (Larsen et al., 2017) high risk for psychosis, no study to date has looked at this in healthy individuals assessed for PLE. We note that the participants included here were in the lower range of the PQ scale. Although we attempted to have a wide spread of psychotic like experiences, participants scoring higher on the PQ scale also scored high on either the Beck anxiety or depression inventory and therefore were not included in the study. The question as to whether the generation of 40 Hz synchrony is related to the degree of psychotic like experiences should be examined in a group with a wider spread of psychotic-like experiences and possibly in a larger sample, as these trait effects are likely small. A scope for future studies is to investigate whether predictability modulates phase locking of the 40 Hz ASSR in people with psychosis as well as how the structure-function relationship is affected.

## Conclusion

The current study introduces a novel combination of 40 Hz ASSR with a classical oddball paradigm. This paradigm introduces the possibility to examine both predictive processes and entrained oscillatory activity in the lower gamma range. In addition, it offers the possibility to study the interaction between the two. We show that the generation of synchronous neural activity in the lower gamma range is linked to the degree of myelination in the right cerebellum. Therefore, our results confirm that the microstructure of the cortex support functional networks.

## Data Availability

All data produced in the present study are available upon reasonable request to the authors

## Acknowledgements

We would like to thank all participants in this study for their time as well as the radiographers at the University of Queensland, Mr. Aiman Al-Najjar and Ms. Nicole Atcheson.

This work was funded by the Australian Research Council Centre of Excellence for Integrative Brain Function (ARC Centre Grant CE140100007). KML received funding from the Lundbeck foundation (R322-2019-2311)

## Supplementary material

**Sup. Figure 1:**
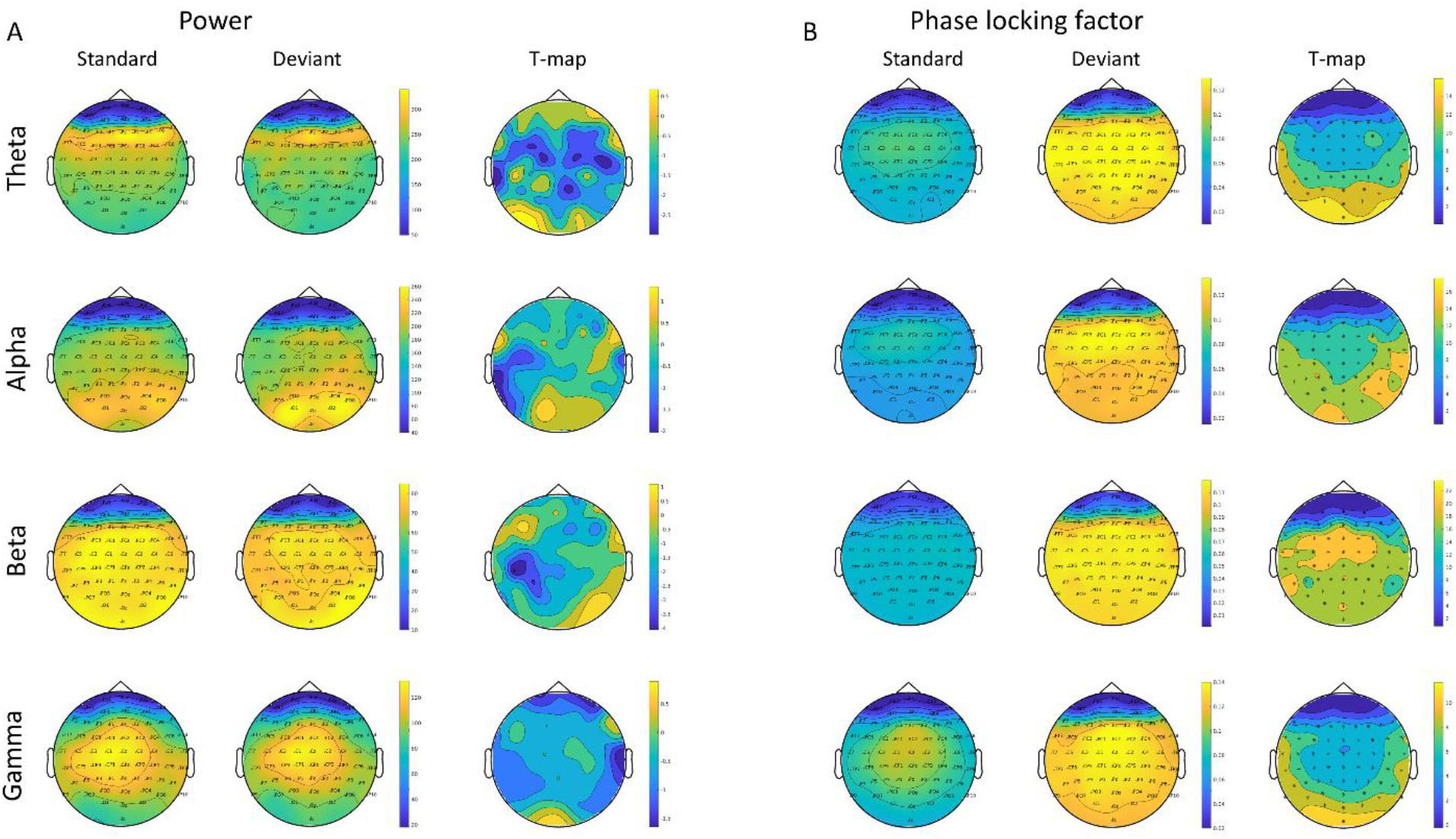
Topographical representation of power (A) and phase-locking factor (B) extracted for the freqneucy bands theta, alpha, beta and gamma. Standard trials are presented in the first column in A and B. Likewise the deviant trials are represented in the second column and finally the statistical T-map in column three, showing results from testing the difference between standard and deviant sounds. In the statistical map, significant electrodes are marked with an Asterix.

**Sup. Figure 2:**
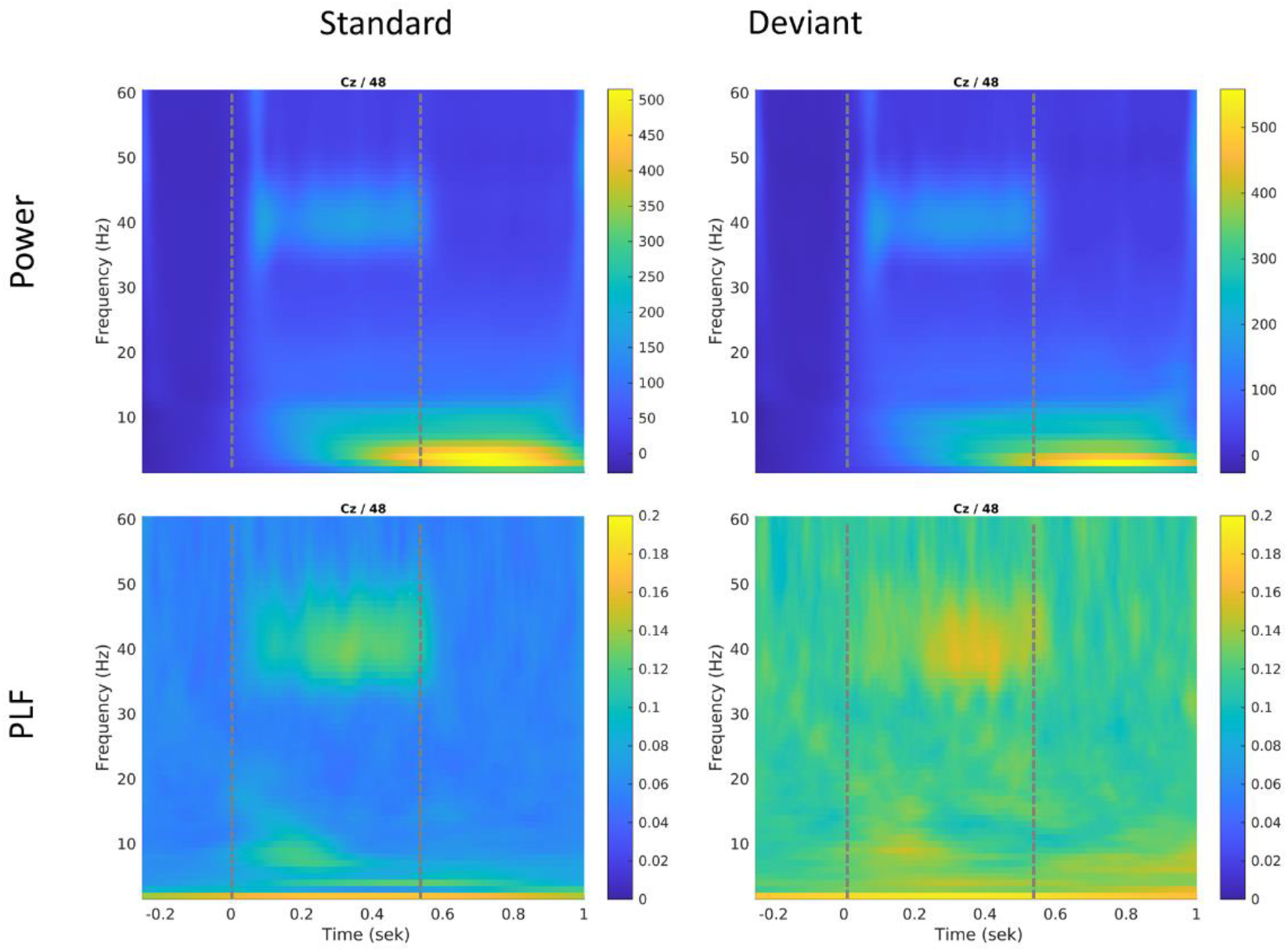
Time-frequency responses at channel Cz (for the short block). Dotted lines indicate onset and offset of auditory stimuli.

